# Real-life application of a point-of-care biosensor for phenylalanine in patients with phenylketonuria

**DOI:** 10.1101/2025.10.01.25336790

**Authors:** Corentin Gondrand, Anna T. Reischl-Hajiabadi, Estelle Bonedeau, Nicolas Weiß, Gerd Schneider, Maurice Fahn, Frederic Myers, Philipp Minel Städle, Kathrin Jeltsch, Georg F. Hoffmann, Jürgen G. Okun, Dorothea Haas, Sven F. Garbade, Kai Johnsson, Thomas Opladen

**Affiliations:** Department of Chemical Biology, Max Planck Institute for Medical Research, Jahnstr. 29, 69120 Heidelberg, Germany; Heidelberg University, Medical Faculty of Heidelberg, Department of Pediatrics I, Division of Pediatric Neurology and Metabolic Medicine, Im Neuenheimer Feld 430, 69120 Heidelberg, Germany; phellow seven GmbH, Bergheimer Str. 147, 69115 Heidelberg, Germany; Frederic Myers Consulting (JERY), Johanniterweg 4, 72768 Reutlingen, Germany; École Polytechnique Fédérale de Lausanne, Institute of Chemical Sciences and Engineering, NCCR in Chemical Biology, CH Building, Station 6, 1015 Lausanne, Switzerland

**Keywords:** PKU, phenylalanine hydroxylase deficiency, point-of-care testing, biosensor

## Abstract

Phenylketonuria (PKU) is a rare metabolic disorder characterized by elevated phenylalanine (PHE) levels, requiring lifelong dietary management and regular monitoring. Current PHE monitoring methods, such as tandem mass spectrometry (FIA-MS/MS), are laboratory-dependent, necessitating sample transport and causing delays in result delivery. This emphasizes the need for point-of-care (PoC) solutions. We developed a bioluminescence-based PHE sensor integrated with the *phenyx* smartphone app for real-time monitoring and data sharing. A clinical study involving 47 PKU patients compared at-home sensor measurements with dried blood spots (DBS) analyzed by FIA-MS/MS. The sensor results were slightly higher than FIA-MS/MS, with a mean difference of 1.4 mg/dL (84.7 µmol/L). Variability in measurements was influenced by test supervision (clinic vs. home) and test kit age (larger differences with older kits, p = 0.001). The sensor and app demonstrated feasibility, analytical reliability, and comparable performance to FIA-MS/MS, providing a potential platform for patient self-testing and improved PHE monitoring.

## Introduction

Phenylketonuria (PKU; OMIM #261600) is one of the most common inherited metabolic diseases with a birth prevalence of approximately 1 in 10,000. It is an autosomal recessive inherited disorder affecting the conversion of the essential amino acid phenylalanine (PHE) to tyrosine due to impaired activity of phenylalanine hydroxylase (PAH; EC 1.14.16.1). Under physiological conditions, PAH converts phenylalanine into tyrosine using the cofactor tetrahydrobiopterin (BH_4_). In PKU, the deficient *PAH* gene leads to accumulation of PHE in blood and all body fluids. PHE is thus a reliable biomarker for diagnosis of PKU, e.g., in newborn screening, but also for lifelong disease monitoring.^1–4^

If untreated, PKU causes severe intellectual impairment and additional neurological symptoms. Therefore, affected patients require lifelong dietary or drug treatment to maintain PHE concentrations within an age-appropriate target range.^2–6^ PHE levels can be measured in dried blood spots (DBS) or plasma by tandem mass spectrometry (FIA-MS/MS), high-performance liquid chromatography, or amino acid analyzer.^4–7^ While these methods are effective, they are time-consuming, expensive, and limited to specialized laboratories. When collecting DBS and sending the DBS cards to the laboratory, the delivery and laboratory turnaround times cause a delay of several days in receiving the test results, highlighting the need for adaptable test formats. Point-of-care (PoC) and at-home tests provide fast results outside traditional laboratory settings. These tests enhance accessibility, affordability, and patient compliance, especially for those requiring frequent monitoring, such as PKU patients.

Although no commercial solution is yet available, numerous studies have successfully developed and described additional quantitative methods for measuring PHE concentrations. These include electrochemical biosensors based on voltammetry, amperometry, potentiometry, or impedimetric, which can be integrated into handheld devices.^8–11^ Optical biosensors including colorimetric approaches such as gold nanoparticles-integrated paper tests^12^ or enzymatic tests using phenylalanine ammonia-lyase (PheCheck^TM^)^13^, as well as fluorimetric methods such as fibre-optic sensors^14^, and luminometric approaches such as chemiluminescent microfluidic assays^15^ or UV-activated smartphone-adapted fluorescent sensors.^16^ Additional solutions include mass-sensitive immunosensors^17^ and surface plasmon resonance chips.^18^

While these technologies show significant promises over the standard of care in terms of simplicity, speed, and cost, the transition from laboratory prototypes to practical, patient-friendly solutions is challenging. Many existing methods are limited by their validation in simplified matrices such as blood serum,^8,9,12,18^ need for specialized equipment,^14,15,18^ or a multiple-step workflow,^16^ restricting their applicability in PoC settings. Beyond the PheCheck^TM^ method,^13^ which is currently being evaluated in a feasibility study with PKU patients (NCT05998109), the performance of other technologies in real-world conditions, particularly in the hands of patients or caregivers at home, remains largely unevaluated.

We had previously developed a bioluminescence-based method enabling PHE quantification at PoC from whole blood ^19^. Results from patient samples were in agreement with those obtained with the standard of care, FIA-MS/MS. Based on this method, we designed a test kit, referred to as PHE sensor, designed for at-home self-testing by PKU patients. We integrated this PHE sensor with a smartphone readout and smartphone health application referred to as *phenyx* app to enable real-time PHE level monitoring and data sharing with healthcare providers. This integration aims to support remote patient monitoring and personalized care, identify trends in PHE levels, flagging issues like poor dietary compliance, medication errors, or fluctuations during illness, and enabling recommendations for adjustments to treatment or diet.

The primary objective of this study was to evaluate the reliability of the PHE sensor and *phenyx* app when used by PKU patients, in a real-word, at-home environment. Secondary objectives included evaluating the differences in duration between blood sampling and results obtained from at-home and FIA-MS/MS, as well as assessing the practicability of the PHE sensor and the *phenyx* app.

## Results

### Study population

All PKU patients interested in participating in the study were included. Between February 2024 and April 2025, 47 patients (N=17 male, N=30 female) with genetically confirmed PKU and a median age of 10.0 years (range 0.7–53.4 years) participated across five cohorts (Table 1). Five test batches were produced with three batches used for group 1, two batches each for groups 2–4, and one batch for group 5. During the study a total of 19 different iPhones were used.

**Table 1:**
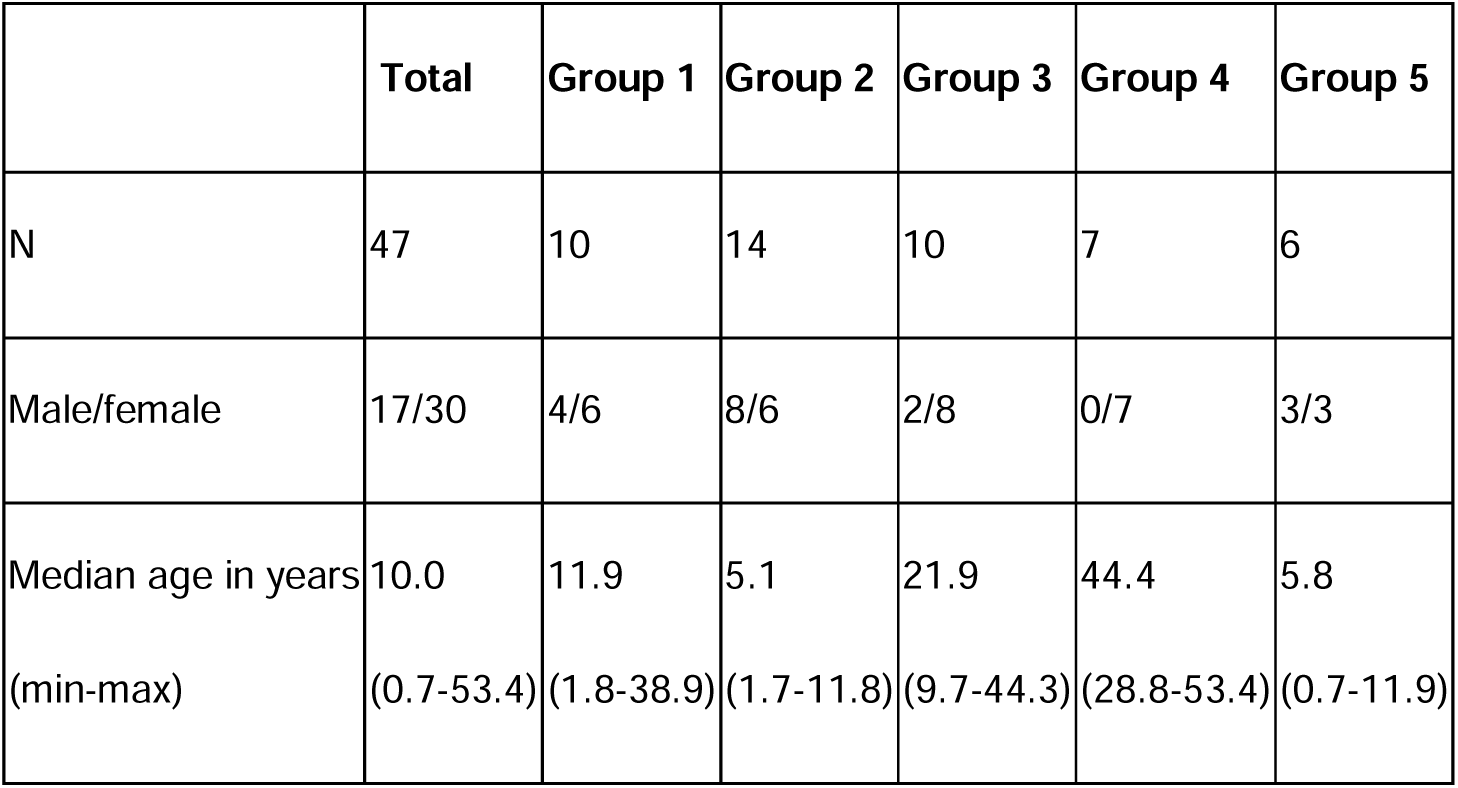
Age and sex distribution of the study groups.

|  | **Total** | **Group 1** | **Group 2** | **Group 3** | **Group 4** | **Group 5** |
| --- | --- | --- | --- | --- | --- | --- |
| N | 47 | 10 | 14 | 10 | 7 | 6 |
| Male/female | 17/30 | 4/6 | 8/6 | 2/8 | 0/7 | 3/3 |
| Median age in years | 10.0 | 11.9 | 5.1 | 21.9 | 44.4 | 5.8 |
| (min-max) | (0.7-53.4) | (1.8-38.9) | (1.7-11.8) | (9.7-44.3) | (28.8-53.4) | (0.7-11.9) |

A total of 194 parallel measurements using the PHE sensor and DBS (FIA-MS/MS) were included in the analysis: study group 1: N=45; group 2: N=55; group 3: N=44; groups 4 and 5: N=25 each (Supplementary Figure S1). When sub-grouped according to age-dependent target ranges, N=116 were in the <6□mg/dl range, N=10 between 6 and 10□mg/dl, N=14 between 10 and 15□mg/dl, and N=54 between 15 and 20□mg/dl. N=50 measurement pairs were excluded due to out-of-range PHE sensor values (>33 mg/dl), the use of plasma instead of DBS, PHE sensor mishandling, improper kit storage and DBS transport issues (Supplementary Table S1).

### Comparison of PHE levels from the PHE sensor and DBS (FIA-MS/MS)

Overall, the PHE sensor measured slightly higher than FIA-MS/MS, with an average difference of 1.4 mg/dl (84.7 µmol/l; linear effects model, p *=* 0.0017). When divided into age-dependent target ranges, the mean differences were 1.65 mg/dl (99.8 µmol/l) for the range <6 mg/dl, 2.38 mg/dl (144.1 µmol/l) for 6–10 mg/dl, –0.32 mg/dl (19.3 µmol/) for 10–15 mg/dl, and 0.69 mg/dl (41.7 µmol/l) for 15–20 mg/dl. Large differences between PHE sensor and DBS FIA-MS/MS measurements were a result of extreme PHE sensor measurements (Figure 1 and Supplementary Figure S2).

**Figure 1:**
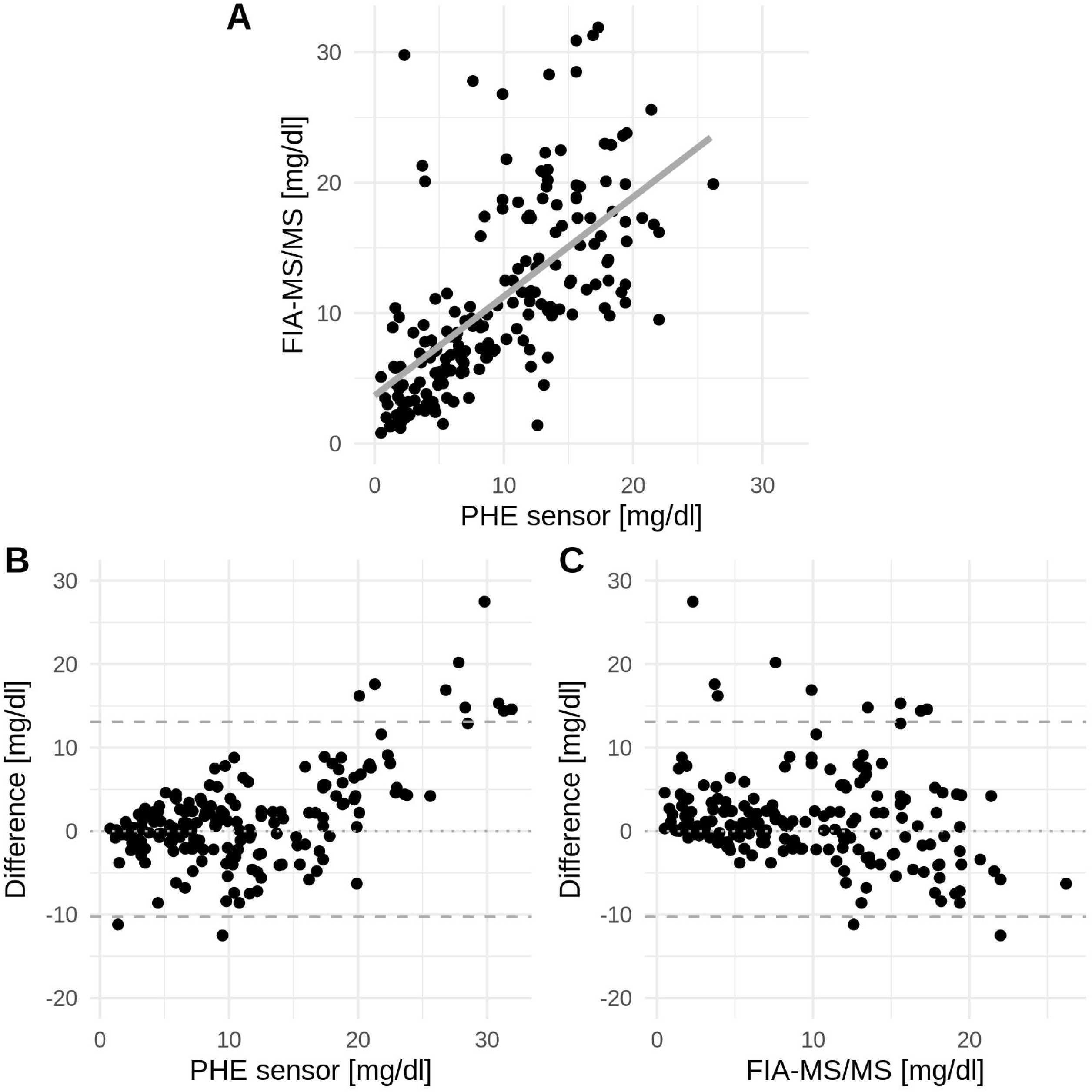
A: Scatterplot with of PHE sensor and DBS FIA-MS/MS measurements. Gray solid lines indicate estimated regression slope from LME model. B and C: Difference of PHE sensor and DBS over PHE sensor measurements (B) and DBS measurements (C). Upper and lower dashed lines are upper and lower tolerance limits. It can be concluded that extreme differences between PHE sensor and DBS measurements are a result of extreme PHE sensor measurements compared to corresponding DBS measurements. The x- and y-axes are labeled from 0 to 30 mg/dl PHE (0 to 1,800 µmol/l) and from –20 to 30 mg/dl (–1,200 to 1,800 µmol/l).

Measurement differences increased with the PHE sensor batch age: the older the test batch, the greater the difference between the measurements (β_batch age_ = 0.0620; p = 0.001, LME) (Figure 2). The mean batch age of differences outside the upper tolerance limit was 77 days (median: 80 days).

**Figure 2:**
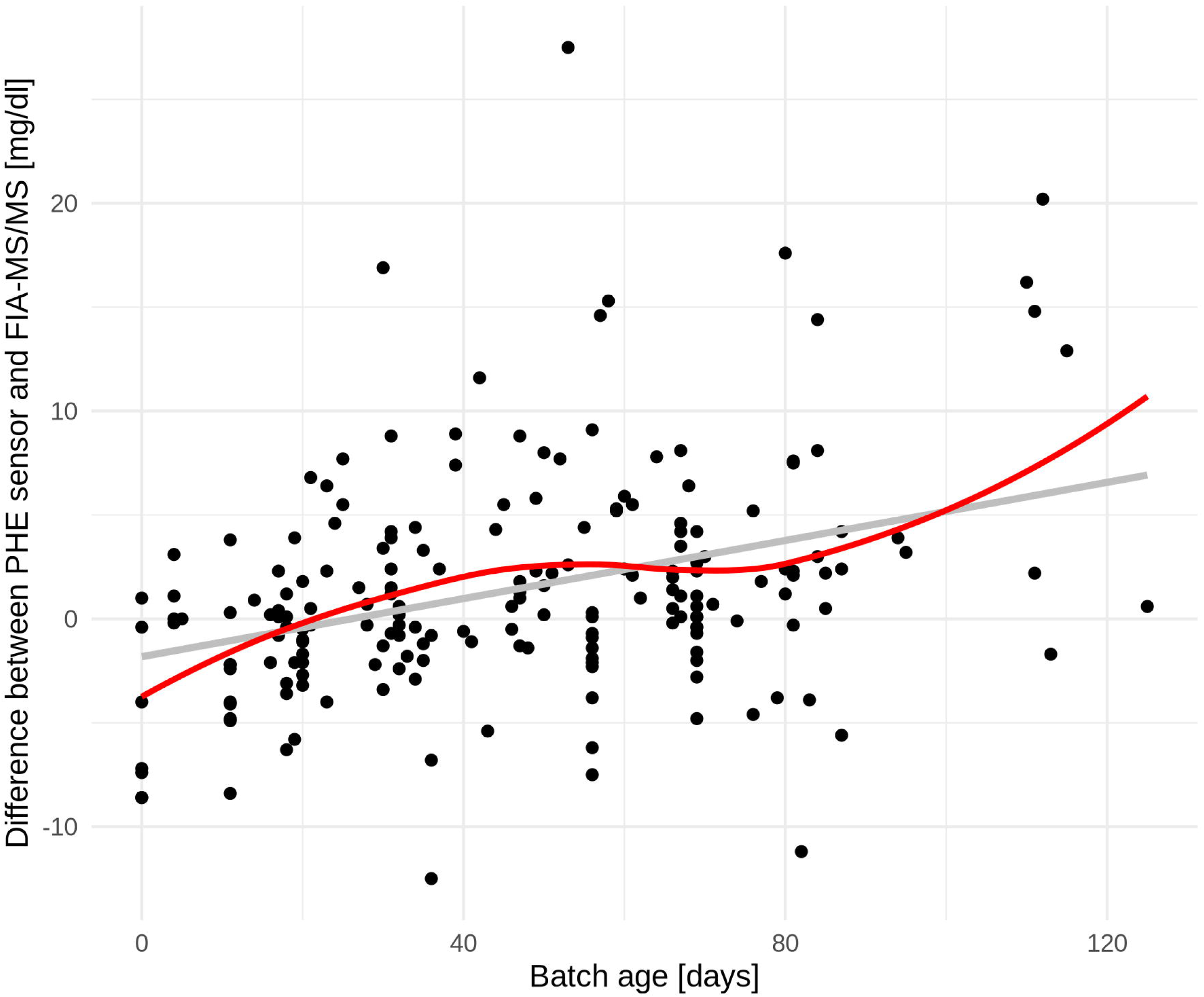
Difference of PHE sensor and DBS across age of batch. Regression line was estimated in LME model, solid red line shows a scatterplot smoothing function. The y-axis is labeled from –10 to 20 mg/dl PHE (–600 to 1,200 µmol/l).

At-home measurements showed a larger variation (mean 2.42 mg/dl) compared to those from the first supervised test (mean −1.88 mg/dl) (p = 0.0016). Batches used in supervised tests were newer (mean 35 days) compared to at-home tests (mean 50 days, p < 0.0001). After adjusting for batch age, supervised PHE sensor measurements showed significantly lower differences from DBS measurements than at-home measurements (LME: p = 0.0001).

The differences between the devices are not significant (LME: p = 0.2018). Also, the age of the participants, sex, batch number, infection status and confidence in handling rated in the questionnaire have no significant impact on differences of the measurements (Table 2).

**Table 2:**
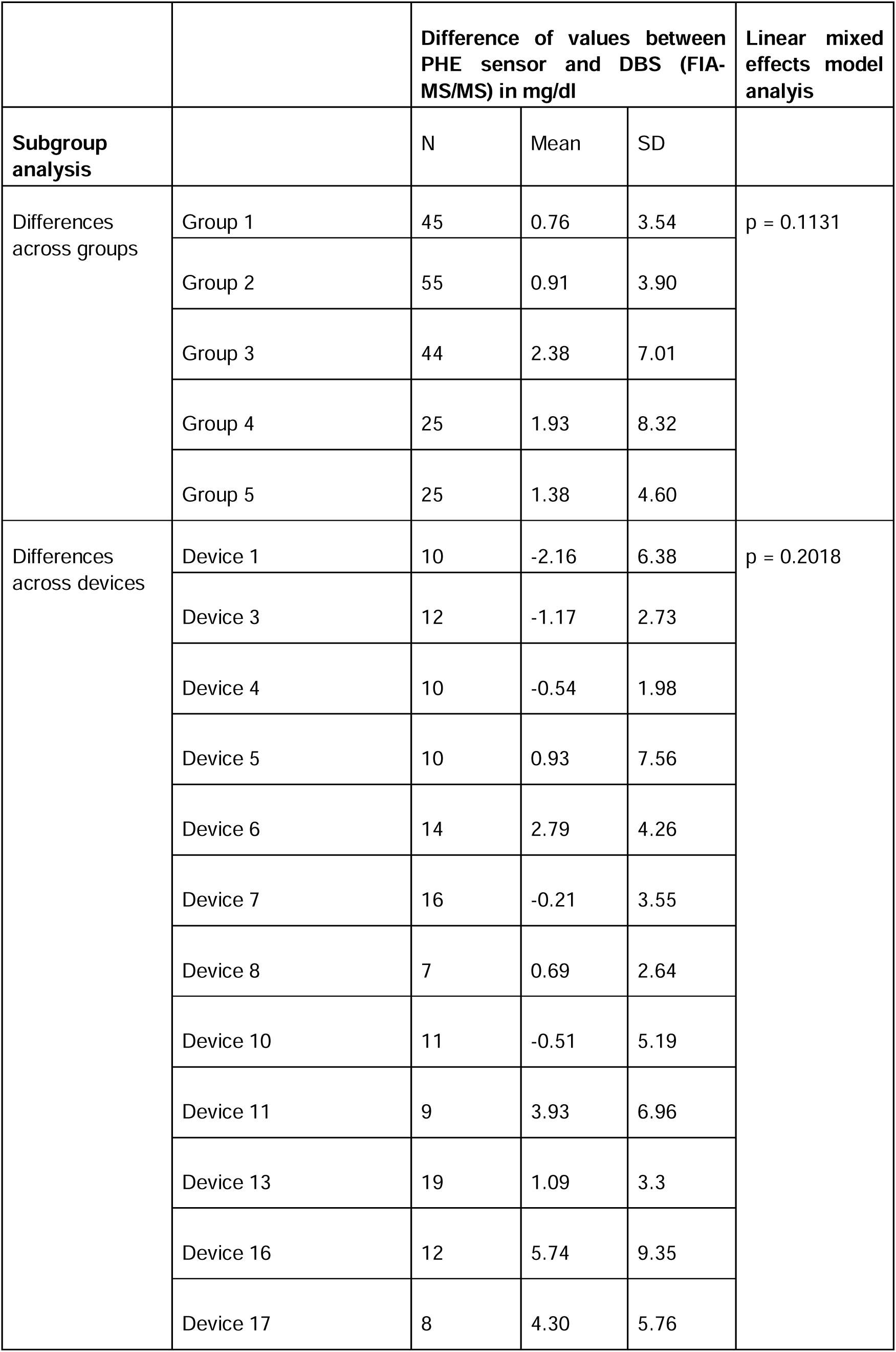
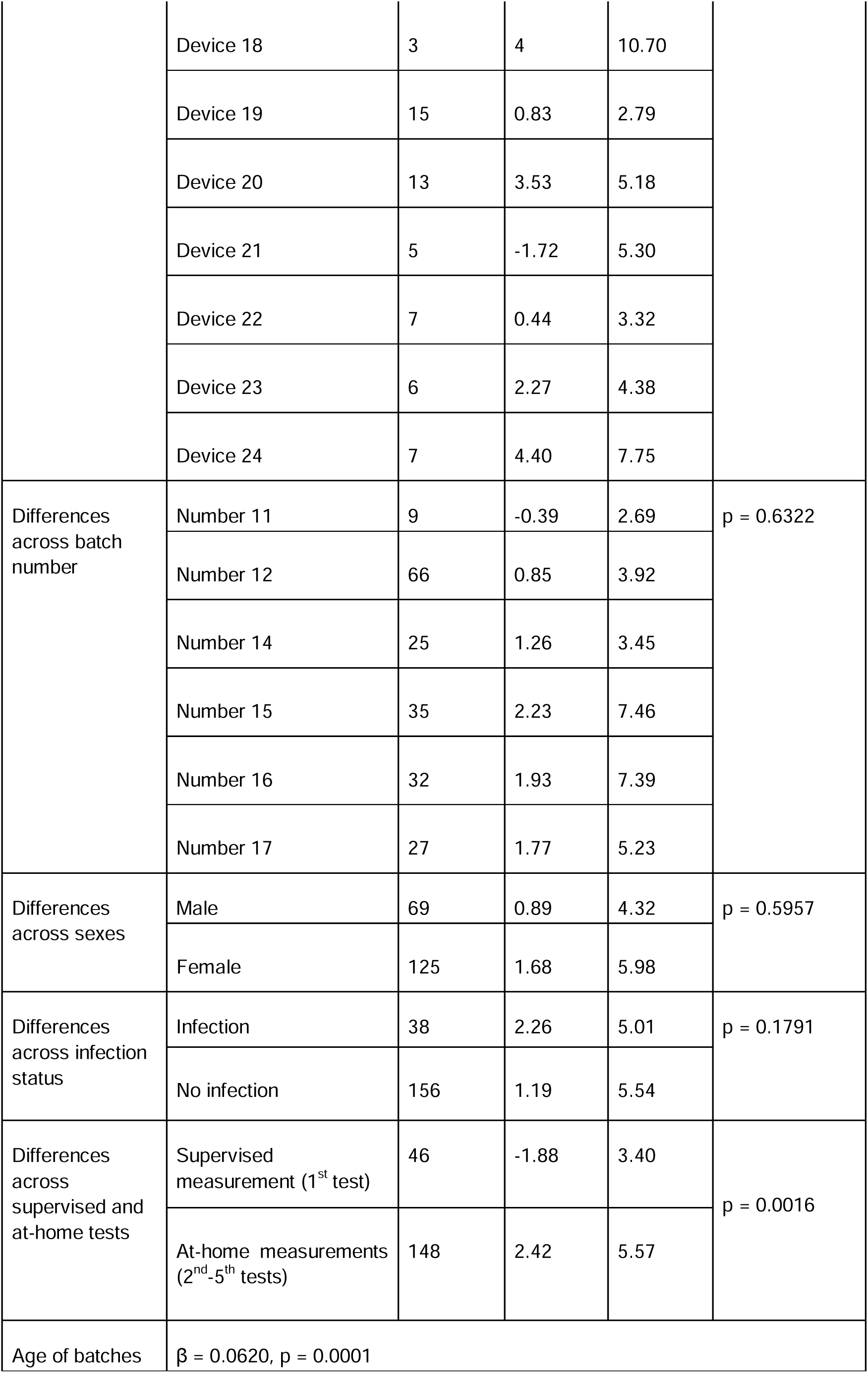
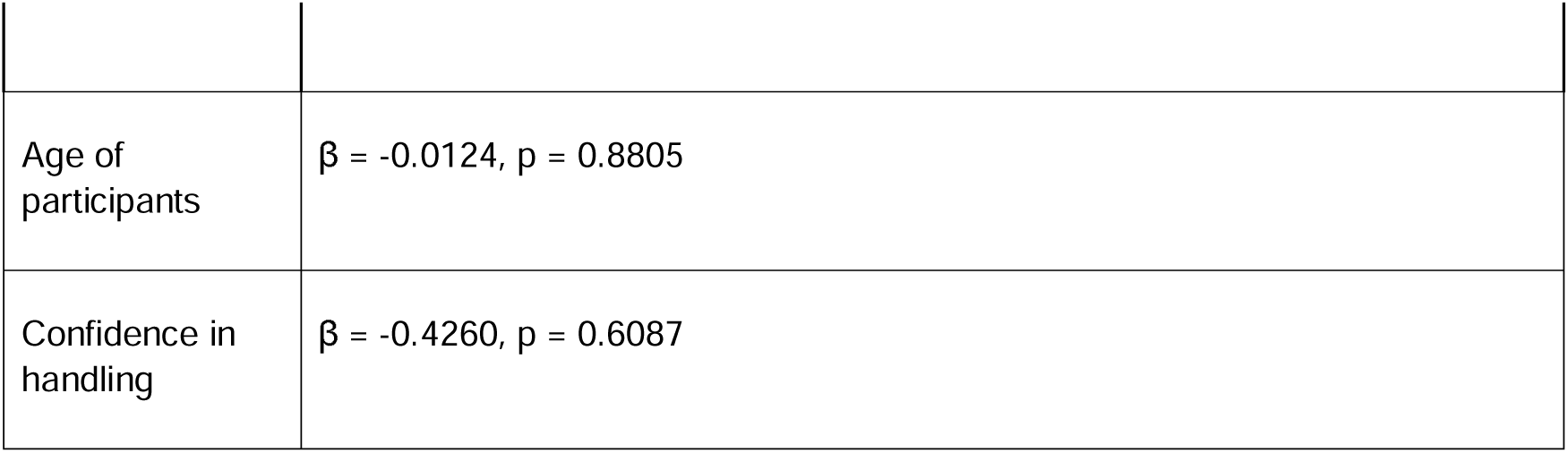
Results from linear mixed effect models.

|  |  | Difference of values between PHE sensor and DBS (FIA-MS/MS) in mg/dl |  |  | Linear mixed effects model analysis |
| --- | --- | --- | --- | --- | --- |
| Subgroup analysis |  | N | Mean | SD |  |
| Differences across groups | Group 1 | 45 | 0.76 | 3.54 | p = 0.1131 |
|  | Group 2 | 55 | 0.91 | 3.90 |  |
|  | Group 3 | 44 | 2.38 | 7.01 |  |
|  | Group 4 | 25 | 1.93 | 8.32 |  |
|  | Group 5 | 25 | 1.38 | 4.60 |  |
| Differences across devices | Device 1 | 10 | -2.16 | 6.38 | p = 0.2018 |
|  | Device 3 | 12 | -1.17 | 2.73 |  |
|  | Device 4 | 10 | -0.54 | 1.98 |  |
|  | Device 5 | 10 | 0.93 | 7.56 |  |
|  | Device 6 | 14 | 2.79 | 4.26 |  |
|  | Device 7 | 16 | -0.21 | 3.55 |  |
|  | Device 8 | 7 | 0.69 | 2.64 |  |
|  | Device 10 | 11 | -0.51 | 5.19 |  |
|  | Device 11 | 9 | 3.93 | 6.96 |  |
|  | Device 13 | 19 | 1.09 | 3.3 |  |
|  | Device 16 | 12 | 5.74 | 9.35 |  |
|  | Device 17 | 8 | 4.30 | 5.76 |  |
|  | Device 18 | 3 | 4 | 10.70 |  |
|  | Device 19 | 15 | 0.83 | 2.79 |  |
|  | Device 20 | 13 | 3.53 | 5.18 |  |
|  | Device 21 | 5 | -1.72 | 5.30 |  |
|  | Device 22 | 7 | 0.44 | 3.32 |  |
|  | Device 23 | 6 | 2.27 | 4.38 |  |
|  | Device 24 | 7 | 4.40 | 7.75 |  |
| Differences across batch number | Number 11 | 9 | -0.39 | 2.69 | p = 0.6322 |
|  | Number 12 | 66 | 0.85 | 3.92 |  |
|  | Number 14 | 25 | 1.26 | 3.45 |  |
|  | Number 15 | 35 | 2.23 | 7.46 |  |
|  | Number 16 | 32 | 1.93 | 7.39 |  |
|  | Number 17 | 27 | 1.77 | 5.23 |  |
| Differences across sexes | Male | 69 | 0.89 | 4.32 | p = 0.5957 |
|  | Female | 125 | 1.68 | 5.98 |  |
| Differences across infection status | Infection | 38 | 2.26 | 5.01 | p = 0.1791 |
|  | No infection | 156 | 1.19 | 5.54 |  |
| Differences across supervised and at-home tests | Supervised measurement (1<sup>st</sup> test) | 46 | -1.88 | 3.40 | p = 0.0016 |
|  | At-home measurements (2<sup>nd</sup>-5<sup>th</sup> tests) | 148 | 2.42 | 5.57 |  |
| Age of batches | β = 0.0620, p = 0.0001 |  |  |  |  |

|  |  |
| --- | --- |
| Age of participants | $\beta = -0.0124, p = 0.8805$ |
| Confidence in handling | $\beta = -0.4260, p = 0.6087$ |

In 37 instances, FIA-MS/MS values were within the age-related target range, whereas the PHE sensor was not (Table 3). In these cases, the mean difference was 8.19 mg/dl (495.8 µmol/l), and the mean batch age was 62.1 days. There were significantly more false positive cases (43.02%), in which the PHE sensor value was outside the target range while the DBS value was within range, than false negatives cases (15.74%) in which the opposite was true (McNemar’s χ² test with continuity correction: p = 0.009722). When analyzing only samples with a batch age below 77 days (165 measurements), the false positive rate reduced to 36.23%, with a mean difference of 7.08 mg/dl (428.6 µmol/l). The overall agreement between the two methods in classifying patients correctly into target vs. out-of-target range was 72.2%. This aligns with the observed positive bias of the PHE sensor test, making it more likely to classify patients as above their target range.

**Table 3:** False-positive measurements with the PHE sensor. The table shows the 37 cases in which PHE sensor measurements were above the age-dependent target range, while the corresponding FIA-MS/MS measurements were within the target range.

| PHE sensor [mg/dl] | DBS FIA-MS/MS [mg/dl] | Difference in PHE value [mg/dl] | Age-dependent target range [mg/dl] | Batch age [days] |
| --- | --- | --- | --- | --- |
| 20.2 | 13.4 | 6.8 | <20 | 21 |
| 23 | 17.8 | 5.2 | <20 | 76 |
| 20.8 | 13.1 | 7.7 | <20 | 25 |
| 20.9 | 12.9 | 8 | <15 | 50 |
| 22.5 | 14.4 | 8.1 | <15 | 84 |
| 8.6 | 5.6 | 3 | <6 | 70 |
| 8.9 | 1.4 | 7.5 | <6 | 81 |
| 10.4 | 1.6 | 8.8 | <6 | 31 |
| 8.2 | 5.9 | 2.3 | <6 | 17 |
| 6.6 | 4.3 | 2.3 | <6 | 66 |
| 7.1 | 4.7 | 2.4 | <6 | 80 |
| 6.9 | 3.5 | 3.4 | <6 | 30 |
| 7.8 | 3.9 | 3.9 | <6 | 31 |
| 7.2 | 4.8 | 2.4 | <6 | 31 |
| 7.9 | 4.4 | 3.5 | <6 | 67 |
| 6.7 | 4.3 | 2.4 | <6 | 87 |
| 20.1 | 3.9 | 16.2 | <6 | 110 |
| 29.8 | 2.3 | 27.5 | <6 | 53 |
| 9.1 | 3.8 | 5.3 | <6 | 59 |
| 11.1 | 4.7 | 6.4 | <6 | 68 |
| 6.8 | 5.9 | 0.9 | <6 | 14 |
| 11.5 | 5.6 | 5.9 | <6 | 60 |
| 21 | 13.4 | 7.6 | <20 | 81 |
| 27.8 | 7.6 | 20.2 | <20 | 112 |
| 21.8 | 10.2 | 11.6 | <20 | 42 |
| 23.8 | 19.5 | 4.3 | <20 | 44 |
| 31.9 | 17.3 | 14.6 | <20 | 57 |
| 28.5 | 15.6 | 12.9 | <20 | 115 |
| 30.9 | 15.6 | 15.3 | <20 | 58 |
| 23.6 | 19.2 | 4.4 | <20 | 55 |
| 20.1 | 17.9 | 2.2 | <20 | 111 |
| 26.8 | 9.9 | 16.9 | <20 | 30 |
| 31.3 | 16.9 | 14.4 | <20 | 84 |
| 28.3 | 13.5 | 14.8 | <20 | 111 |
| 6.5 | 5.5 | 1 | <6 | 47 |
| 21.3 | 3.7 | 17.6 | <6 | 80 |
| 8.5 | 3 | 5.5 | <6 | 61 |

### Processing time: PHE sensor versus DBS

The measurement time with the PHE sensor was approximately 15 minutes. This would also be the time it would take for the result to be available to the patient in the future. In comparison, DBS cards from at-home measurements arrived at the laboratory after a median time of 2 days (range 0–11 days), and patients received the results after a median of 3 days (range 1–11 days) after the measurement date.

### Patient questionnaire: Feasibility of the PHE sensor and *phenyx* app

Of the 47 participants, 46 (98%) completed the questionnaire (36% patients’ mothers, 30% female adult patients, 12% fathers, 8% boys, 6% parents, 6% girls, 2% male adult patients).

Instructions during onboarding and the user manual were rated as “very good” (87%) and “good” (13%). Confidence during measurement was reported as “very confident” (37%), “confident” (46%), “mostly confident” (15%), or “okay” (2%). Confidence in handling increased with repeated use: 41% of participants felt confident after the first test, 48% after the second, and 9% after the third. However, 2% never felt fully confident, likely due to the monthly testing interval, suggesting a gradual learning process.

Individual steps of the test procedure were rated “very good” and “good” (median of 1-2). The waiting time until the blood sample was applied received a lower rating (median of 3, 67% “okay”, Supplementary Figure S3). The design of the *phenyx* app was rated “very good” (41%), “good” (52%), and “okay” (7%). Overall handling of the PHE sensor was rated “very good” (56%), “good” (35%), and “satisfactory” (9%). Malfunction issues were reported by 36%, mostly related to the app, with frequent comments on Wi-Fi interruptions.

While 40% rated the PHE sensor as requiring more effort and 4% much more effort than DBS testing (Figure 3), 100% of the participants preferred the PHE sensor over laboratory testing, and 98% would recommend it to other PKU patients.

**Figure 3:**
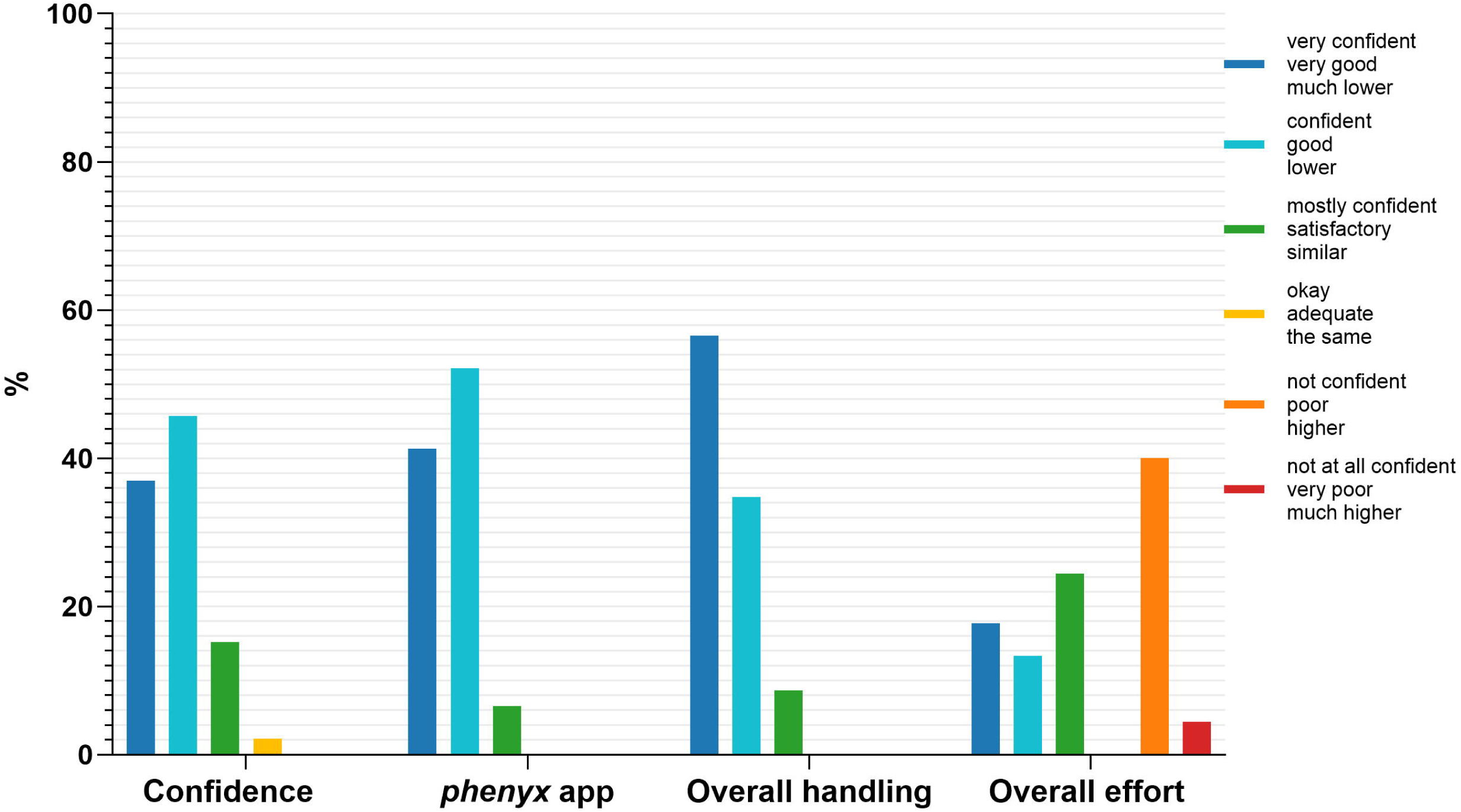
Ratings of confidence during the measurement, design of the *phenyx* app, overall handling of the PHE sensor, and comparison of the overall effort using the PHE sensor and *phenyx* app versus DBS. The bar chart shows the percentage distribution of ratings. N=46 answered the questions on confidence, design of the *phenyx* app and the overall handling. N=45 answered the question on the overall effort.

An LME model showed no significant differences in ratings of confidence during the measurement, overall handling, design of the *phenyx* app, or overall effort among the different groups performing the measurement.

Suggested features for future app development included in-app communication with healthcare providers and dietitians, a food diary and PHE intake tracker, visualization of additional amino acids, and reminders for measurements and appointments.

## Discussion

This is the first clinical study to evaluate PoC measurement of PHE in a home setting in a large cohort of children and adults with PKU. We demonstrate that at-home quantification of PHE using the PHE sensor in combination with the *phenyx* app is feasible, well accepted, and yields results comparable to the current laboratory standard of FIA-MS/MS.

The PHE sensor showed a small positive bias compared to DBS FIA-MS/MS (mean +1.4□mg/dl; 84.7 µmol/l)), which remained within acceptable limits for novel diagnostic technologies. The PHE sensor demonstrated high accuracy across clinically relevant ranges (<6 to 20 mg/dl; <360 to 1200 µmol/l), which is essential for PKU management where adherence to strict age-specific target ranges is critical.^2–6^ Importantly, there was a 72.2% agreement between the PHE sensor and FIA-MS/MS when classifying patients as within or outside their target range, further supporting the sensor’s clinical utility.

Unlike other biosensors that can only detect PHE levels up to approximately 7.2 mg/dl (435.8 µmol/l),^13^ the PHE sensor is able to quantify higher values relevant for older PKU patients. While enzyme-based biosensors using PAH may be affected in PKU patients undergoing Pegvaliase therapy,^1,13^ our NADPH-based PHE sensor should remain unaffected.

A major clinical advantage of our home-based PHE sensor testing is the rapid availability of results: 15 minutes, compared to the median of 3 days for DBS results. This immediate feedback could enable timely dietary adjustments, which are particularly important for children and pregnant women, helping minimize unnecessary dietary restrictions and reduce psychosocial burden.^1^ User acceptance was high, with most participants finding the PHE sensor and *phenyx* app easy to use and preferring them over laboratory testing, despite reporting a higher effort and a learning curve.

Several limitations must be acknowledged. Our study included participants from all age groups and approximately 10% of the PKU patients treated at our center, but the overall sample size and observation period for validating a new device were modest. Self-selection bias may also have favored more motivated users.

Another limitation relates to the reagent stability of the PHE sensor. Greater variability was observed with increasing batch age, with a functional shelf-life calculated at 77 days under real-life conditions. Test refrigeration and regular kit supply posed additional challenges, particularly during travel or in warmer climates. The stability of tube 2, containing lyophilized reagents, may have been affected by moisture absorption before sealing, manual sealing at ambient conditions, and inconsistencies in the lyophilization process. The lyophilizer also lacked integrated freezing and cycle logging, limiting process control. These factors likely contributed to increased batch variability and accelerated reagent degradation. Addressing these technical issues will require improved process control, optimized formulations, automated sealing in controlled environments, and comprehensive stability testing under different humidity and temperature conditions.

Greater variability was observed in at home compared to supervision testing. This could be attributed to user-dependent factors such as incomplete capillary blood collection, insufficient mixing of reagents, or incomplete sample transfer. Smartphone positioning or handling errors might have also contributed to the variability, especially when combined with older batches or suboptimal storage. These findings suggest that extended patient training, a testing schedule with more frequent tests, as well as a simplified, more user-friendly test design, with limited user influence, could help minimize variability in at-home settings.

In the current format, the PHE sensor relies on the integrated iPhone 14 Pro camera and an active internet connection. For broader accessibility, future versions should enable offline measurements and compatibility with a wide range of smartphones. Furthermore, the test should be more automatized. Economic considerations will also be critical as reimbursement and costs have traditionally limited the adoption of new medical devices in rare and pediatric diseases.^20^ However, integrating the system with patients’ existing smartphones could provide a cost-effective solution.

In conclusion, our findings support the use of the PHE sensor and *phenyx* app as a reliable and user-friendly tool for home-based PHE monitoring. Real-world validation demonstrates that this integrated solution has the potential to enhance patient autonomy, improve metabolic control, and reduce both the clinical and psychological burden of PKU. Future studies should investigate long-term use and integration into routine care pathways. Broader implementation could meaningfully transform PKU management by decentralizing care without compromising diagnostic quality. The further development and planned release of the app could facilitate communication between patients and healthcare providers, in our increasingly digital world. Additionally, the *phenyx* app and the PHE sensor kit may be expandable to other inherited metabolic diseases with defined biomarkers, such as maple syrup urine disease, also requiring frequent laboratory measurements and communication between patients and healthcare providers for therapy adjustments.

## Methods

### Study design

This single-center study (German Clinical Trials Register; trial registration ID: DRKS00031972) was conducted at Heidelberg University Hospital from February 2024 to April 2025 and approved by the local ethics committee (University Hospital Heidelberg, Germany; application number: S-109/2023). The study was open to all interested patients with genetically confirmed PKU. The study followed routine PHE monitoring as recommended by the German consensus guidelines for PKU (updated version 2025 under review, AWMF 027 - 002), no additional interventions were necessary. Written informed consent was obtained from all participants or their legal guardians. None of the patients were treated with Pegvaliase and one pregnant woman received sapropterin.

PHE concentrations were measured in capillary blood at PoC using the PHE sensor, with parallel measurements in DBS analyzed by FIA-MS/MS at five predefined time points. The parallel measurements required only 5 µl of additional blood without extra pricks. The study adhered to the routine monitoring schedule, which recommends biweekly PHE monitoring for patients aged 0–12 years, and monthly monitoring for patients aged 13 years and older. This resulted in a study duration of approximately 2 and 4 months, respectively. Following an initial onboarding appointment at the outpatient clinic, including training and first supervised parallel measurements, the remaining four measurements were conducted independently at home (Supplementary Table S2). The study was conducted sequentially in five cohorts. The date of receipt of DBS in the laboratory and the date of reporting the results were obtained from the laboratory information system.

### Questionnaire

At the end of the study, participants or caregivers completed a 23-item questionnaire regarding their user experience of the PHE sensor and the *phenyx* app. The survey covered usability, handling issues, confidence in handling, perceived effort compared to laboratory testing, preference for PHE sensor use, and desired app features (Supplementary Table S3).

### PHE sensor

The PHE sensor was adapted from a bioluminescence-based PHE PoC assay ^19^ developed at the Max Planck Institute for Medical Research. In this method, PHE is converted by an NADP^+^-dependent phenylalanine dehydrogenase into NADPH, which is quantified by a bioluminescent NADPH sensor (Supplementary Figure S4). The PHE sensor is a test kit containing a lancet, two single-use pipettes, a tube containing buffer (tube 1), a tube with lyophilized reagents (tube 2), a paper test with four detection zones, and a smartphone holder (Figure 4). The *phenyx* app guided users through the test steps (summary of instructions in the Supplementary).

**Figure 4:**
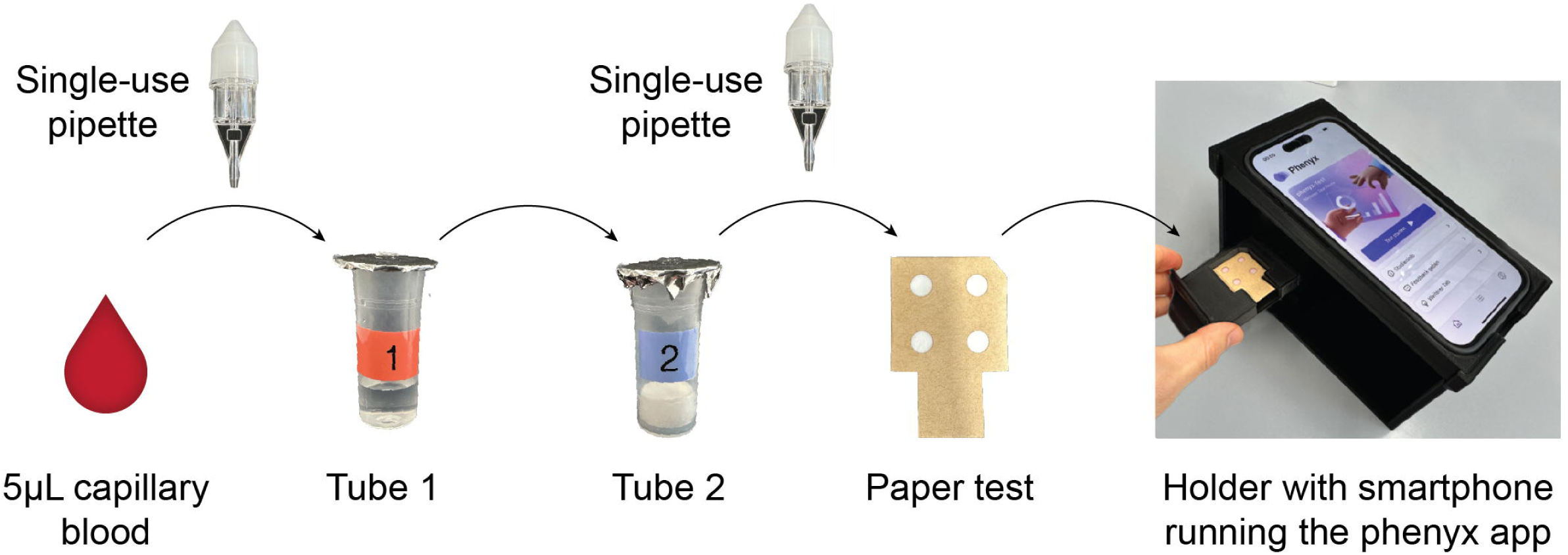
PHE sensor measurement process. A volume of 5 μL of blood obtained from a finger prick is collected with a single-use pipette. The blood-filled pipette is clicked onto tube 1. The content of tube 1 is then transferred into tube 2, followed by a 5-minute incubation. A drop from tube 2 is then applied on each detection zone of the paper test, resulting in the light emission of the NADPH sensor. The paper test is inserted into the drawer of the smartphone holder, where a smartphone running the *phenyx* app takes and analyses pictures of the test signal, calculates PHE concentration and transmits the result to the medical team.

Each smartphone used in the study was evaluated with the PHE sensor and results were compared to a reference iPhone 14 Pro to confirm color detection accuracy (Supplementary Figure S5).

Reagents (tube 2 and test paper) were produced in batches of 100-150 units, each batch was calibrated using spiked and PKU patient whole blood samples and the shelf-life was shown to be at least 6 weeks at 4°C (Supplementary Figure S6). For patients on biweekly testing schedules, all kits were supplied at once. Patients tested monthly received two kits during clinic visits, with subsequent tests shipped separately, which introduced logistical constraints. In addition, replacement kits occasionally had to be shipped at short notice (e.g., after handling issues), requiring the redistribution of older batches.

### Digital health application *phenyx* app

The study partner phellow seven GmbH developed the *phenyx* app (<u>phenyx.de</u>) for Apple iOS. In addition to the app for patients, a browser-based application was developed for the supporting team of medical professionals, enabling real-time assessment of patient input and lab results. All participants received an iPhone 14 Pro with the *phenyx* app pre-installed, pre-configured and a unique onboarding QR code. The app guided patients step by step through the PHE sensor measurement process using videos.

For the PHE level quantification, images were taken with the iPhone 14 Pro main camera (exposure 1s, white balance 8,000K, focus distance 0.0, ISO 5,000). Nine images were taken at five-second intervals, beginning 3:45 min after applying the sample to the paper test. Images were transferred through a secure channel to the server, where they were processed and averaged to calculate PHE concentration. If any error occurred during processing (e.g., quality check fails) the system would display an error message to the patient.

The quantification algorithm, adapted from the literature, recognizes the test spots, determines mean pixel intensities in blue and red channels, and calculates their ratio.^21^ PHE level is calculated from this ratio in combination with the calibration data stored in the QR code of each test.

During the course of study, the PHE sensor results were not shared directly with patients to prevent self-directed dietary changes prior to formal evaluation. By contrast, DBS results from FIA-MS/MS were manually uploaded by the study team into the app and displayed as trends together with age-specific target ranges for German-speaking countries (<6 mg/dl (<360 µmol/l) for <12 years and pregnant women; <10 mg/dl (600 µmol/l) for <15 years, <15 mg/dl (900 µmol/l) for <18 years, <20 mg/dl (1200 µmol/l) for >18 years) (Figure 5).

**Figure 5:**
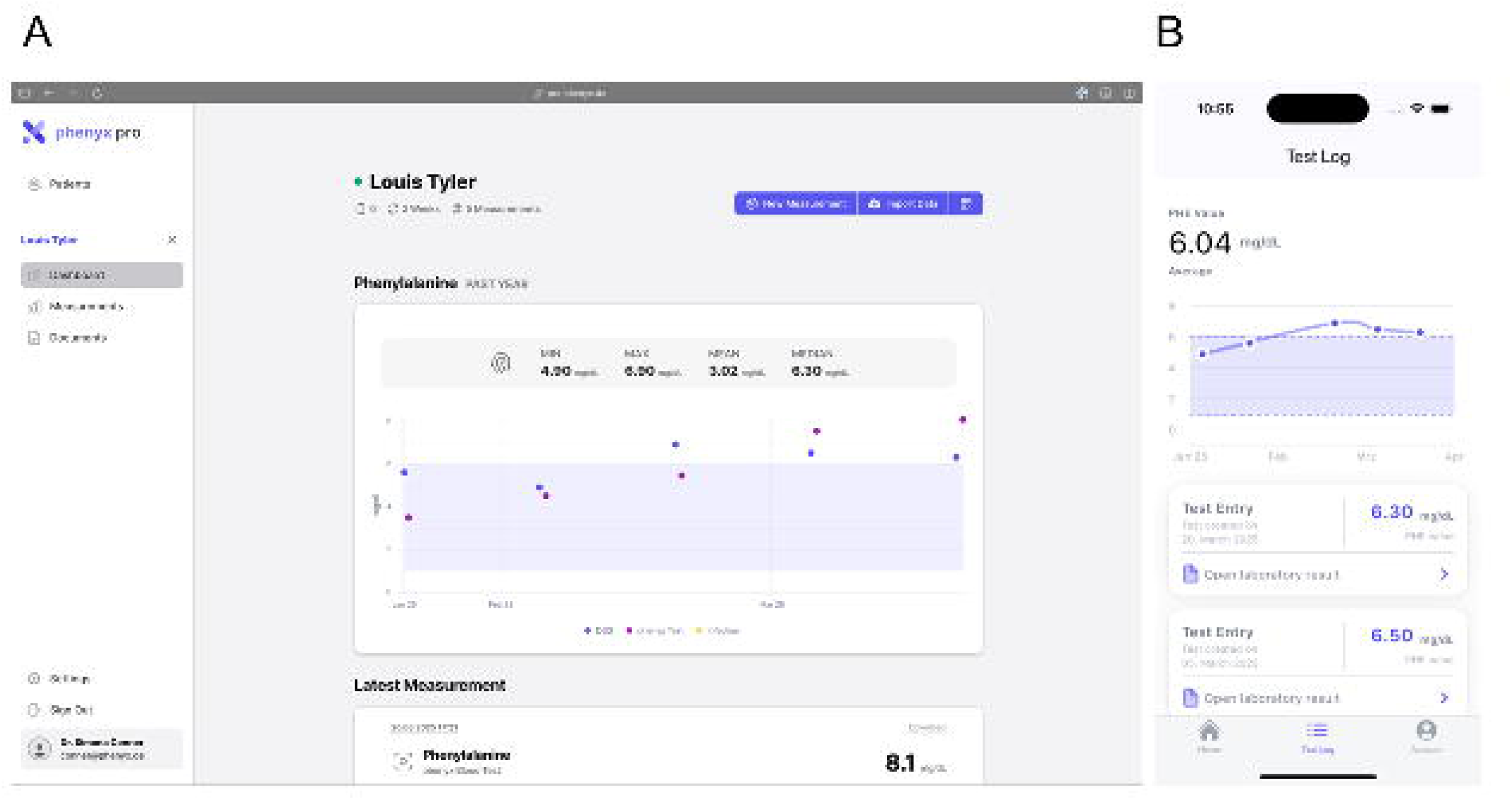
*phenyx* app dashboard. A: Display of the *phenyx* app dashboard from the healthcare provider’s perspective, presenting the PHE values from both the PHE sensor and DBS measurements, along with information on infection status and the patient’s age-specific reference range. B: Display of the *phenyx* app dashboard from the patient’s perspective, showing individual measurements, target range, as well as trends and averages. In the study, only the results from the DBS measurements were shown to the patients. The y-axis is labeled from 0 to 8 mg/dl PHE (0 to 484 µmol/l). Please note that this information refers to a fictional patient and does not contain identifiable data from the study.

### DBS PHE quantification with FIA-MS/MS

DBS samples were measured in the laboratory using the gold standard, FIA-MS/MS.^4–7^ Because PHE concentrations in erythrocytes are 19% lower than in plasma, values were corrected using the slope of the 75th percentile from the quantile regression, as previously described.^7^

### Statistical analysis

Data were analyzed using descriptive and inferential statistical methods. All statistical analyses were conducted using R Version 4.5.0 and GraphPad Prism Version 10 and p-values <0.05 were considered statistically significant.

Exclusion of data from analyses was done if only one measurement (PHE sensor or DBS) was available for a given time point, kits had not been stored properly, plasma rather than DBS was used, testing was delayed for more than 2 weeks, or PHE sensor values were <1 or >33 mg/dl (outside the quantifiable range of the PHE sensor).

The difference between the PHE sensor and FIA-MS/MS was analyzed using a linear mixed-effects (LME) model with a random intercept for the participants’ IDs. The following predictor variables were analyzed in LME: group (study group 1 to 5), device (19 different devices), age (numeric), sex (female/male), test batch (six different batches), test batch age (numeric), test setting (supervised (1st test) versus at-home tests (2nd-5th test) and infection status (yes/no). Because of missing questionnaire data, the effect of the ranking of confidence during measurement (1-6) answered in the questionnaire was evaluated with all predictor variables listed above, but with a lower sample size (N=46, instead of N=47). We used function lme() from R package ‘nlme’ version 3.1-168 to compute LME models. For categorical predictors, we report p-values from analysis of variance tables with marginal sum of squares (function anova() from R package ‘nlme’), otherwise the regression coefficient β and the respective p-value. To compute bias and tolerance limits of measurement differences between PHE sensor and FIA-MS/MS, R package ‘SimplyAgree’, version 0.2.1 was applied.

## Abbreviations

BH_4_: tetrahydrobiopterin
BRET: bioluminescent resonance energy transfer
DBS: dried blood spot
FIA-MS/MS: Flow injection analysis–tandem mass spectrometry
FRET: fluorescence resonance energy transfer
LME: linear mixed-effects model
NADPH: nicotinamide adenine dinucleotide phosphate
PAH: phenylalanine hydroxylase
PHE: phenylalanine
PDH: phenylalanine dehydrogenase
PKU: phenylketonuria
PoC: point-of-care

## Data availability

The data that support the findings of the study are available from the corresponding authors upon reasonable request.

## Code availability

Not applicable.

## Acknowledgements

We gratefully acknowledge M. Lomtadze for his financial support of this study.

We also acknowledge support by the Fraunhofer-Max Planck Cooperation Program and the Consense project funded by the Marie Skłodowska-Curie grant agreement number 955623.

The funder played no role in study design, data collection, analysis and interpretation of data, or the writing of this manuscript.

We acknowledge Daniel Berndt, and Martin Lukat from the Mechanical Workshop at the Max Planck Institute for Medical Research for their assistance with the design and 3D printing of the smartphone holder.

## Author contributions

T.O. and A.T.R.-H. developed and supervised the clinical study. K.J.^1^, C.G. and E.B. conducted the laboratory analyses and the development of the PHE sensor. N.W., G.S. and M.F. developed the *phenyx* app. F.M. and P.M.S. contributed to the design of the *phenyx* app. A.T.R.-H., T.O., C.G., E.B., N.W. and K.J.^2^ contributed to the study design, coordination of the trial, and data collection. S.F.G. performed statistical analyses. D.H. contributed to the data interpretation. D.H. and J.G.O. contributed to laboratory analyses of DBS with FIA-MS/MS. A.T.R.-H., C.G., E.B. and T.O. wrote the first draft of the manuscript. All authors edited or substantively reviewed the publication and had full access to all the data in the study. All authors approved the final version of the manuscript and had responsibility for the decision to submit for publication. The corresponding authors (T.O. and K.J.^1^) had full access to the complete dataset of the study and had final responsibility for the decision to submit for publication.

## References

1. van Spronsen, F.J., et al. Phenylketonuria. Nature reviews Disease primers 7, 36 (2021).

2. Blau, N., Van Spronsen, F.J. & Levy, H.L. Phenylketonuria. The Lancet 376, 1417–1427 (2010).

3. Vockley, J., et al. Phenylalanine hydroxylase deficiency: diagnosis and management guideline. Genet Med 16, 188–200 (2014).

4. Van Wegberg, A., et al. The complete European guidelines on phenylketonuria: diagnosis and treatment. Orphanet journal of rare diseases 12, 1–56 (2017).

5. Singh, R.H., et al. Recommendations for the nutrition management of phenylalanine hydroxylase deficiency. Genetics in Medicine 16, 121–131 (2014).

6. van Spronsen, F.J., et al. Key European guidelines for the diagnosis and management of patients with phenylketonuria. Lancet Diabetes Endocrinol 5, 743–756 (2017).

7. Haas, D., et al. Differences of phenylalanine concentrations in dried blood spots and in plasma: erythrocytes as a neglected component for this observation. Metabolites 11, 680 (2021).

8. Dinu, A. & Apetrei, C. A Review on Electrochemical Sensors and Biosensors Used in Phenylalanine Electroanalysis. Sensors (Basel) 20(2020).

9. Shyam, R., Sekhar Panda, H., Mishra, J., Jyoti Panda, J. & Kour, A. Emerging biosensors in Phenylketonuria. Clinica Chimica Acta 559, 119725 (2024).

10. Li, S., et al. Electrochemical Biosensors for Whole Blood Analysis: Recent Progress, Challenges, and Future Perspectives. Chem Rev 123, 7953–8039 (2023).

11. Parrilla, M., Vanhooydonck, A., Watts, R. & De Wael, K. Wearable wristband-based electrochemical sensor for the detection of phenylalanine in biofluids. Biosens Bioelectron 197, 113764 (2022).

12. Jafari, P., et al. Colorimetric biosensor for phenylalanine detection based on a paper using gold nanoparticles for phenylketonuria diagnosis. Microchemical Journal 163, 105909 (2021).

13. Wada, Y., et al. A method for phenylalanine self-monitoring using phenylalanine ammonia-lyase and a pre-existing portable ammonia detection system. Mol Genet Metab Rep 35, 100970 (2023).

14. Arakawa, T., et al. Biosensor for L-phenylalanine based on the optical detection of NADH using a UV light emitting diode. Microchimica Acta 173, 199–205 (2011).

15. Kamruzzaman, M., et al. Microfluidic chip based chemiluminescence detection of L-phenylalanine in pharmaceutical and soft drinks. Food Chemistry 135, 57–62 (2012).

16. Melman, Y., Katz, E. & Smutok, O. Phenylalanine biosensor based on a nanostructured fiberglass paper support and fluorescent output signal readable with a smartphone. Microchemical Journal 179, 107497 (2022).

17. Kaleli-Can, G., Özgüzar, H.F. & Mutlu, M. Development of QTF-based mass-sensitive immunosensor for phenylketonuria diagnosis. Applied Physics A 128, 277 (2022).

18. Çimen, D., Bereli, N. & Denizli, A. Surface plasmon resonance based on molecularly imprinted polymeric film for L-phenylalanine detection. Biosensors 11, 21 (2021).

19. Yu, Q., et al. Semisynthetic sensor proteins enable metabolic assays at the point of care. Science 361, 1122–1126 (2018).

20. Iqbal, C.W., Wall, J. & Harrison, M.R. Challenges and climate of business environment and resources to support pediatric device development. Seminars in Pediatric Surgery 24, 107–111 (2015).

21. Griss, R., et al. Bioluminescent sensor proteins for point-of-care therapeutic drug monitoring. Nature chemical biology 10, 598–603 (2014).

