## Supplementary for "Real-life application of a point-of-care biosensor for phenylalanine in patients with phenylketonuria"

**Supplementary Material**

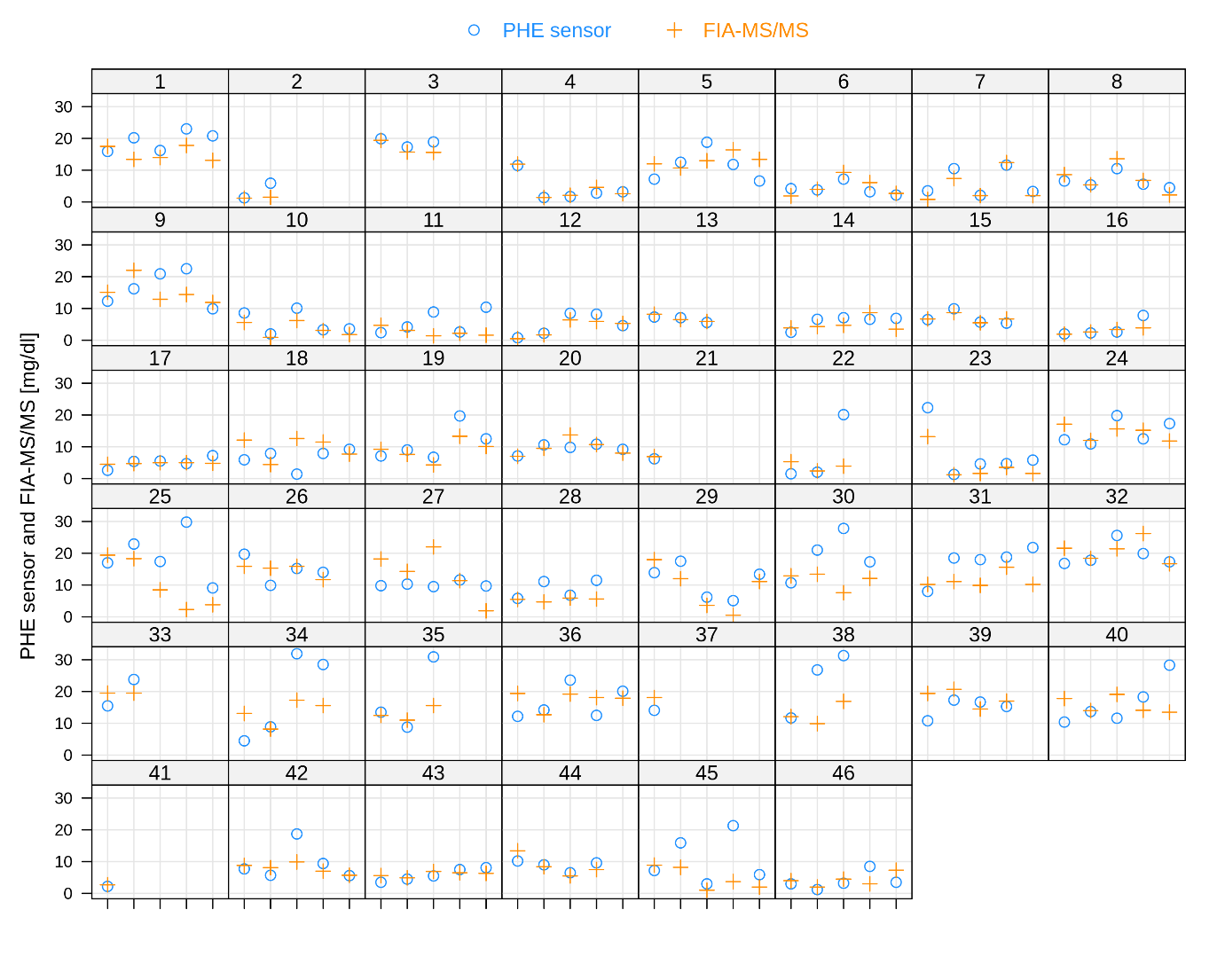

**Figure S1: Individual measurements of PHE sensor and DBS.**

Each square represents one patient, with the panel header indicating patient ID and smartphone device number. The y-axis represents PHE in mg/dl (labeled from 0 to 30 mg/dl, which corresponds to 0 to 1,800 µmol/l), x-axis: date of measurement. Blue circles represent data points obtained with the PHE sensor in combination with the phenyx app. Orange crosses represent data points obtained with DBS measured with FIA-MS/MS.

**Table S1: Exclusion of PHE measurements and reasons.**

| **N** | **Reason for exclusion** |
| --- | --- |
| 12 | PHE value measured with the PHE sensor >33 mg/dl (>1,980 µmol/l); exclusion of PHE sensor and parallel DBS values |
| 3 | Plasma instead of DBS; exclusion of PHE sensor and parallel plasma values |
| 21 | Technical issues with the PHE sensor; no PHE value available from the PHE sensor; exclusion of DBS value |
| 6 | Technical issues with PHE sensor, measurement cancelled and repeated; exclusion of cancelled PHE sensor measurement but maintaining of repeated PHE sensor measurement with parallel DBS measurement |
| 5 | Measurement with PHE sensor kit not stored in fridge; exclusion of PHE sensor and parallel DBS values |
| 1 | DBS card did not arrive in laboratory; exclusion of PHE sensor value |
| 2 | Delayed testing >2 weeks beyond schedule; exclusion of PHE sensor and parallel DBS values |

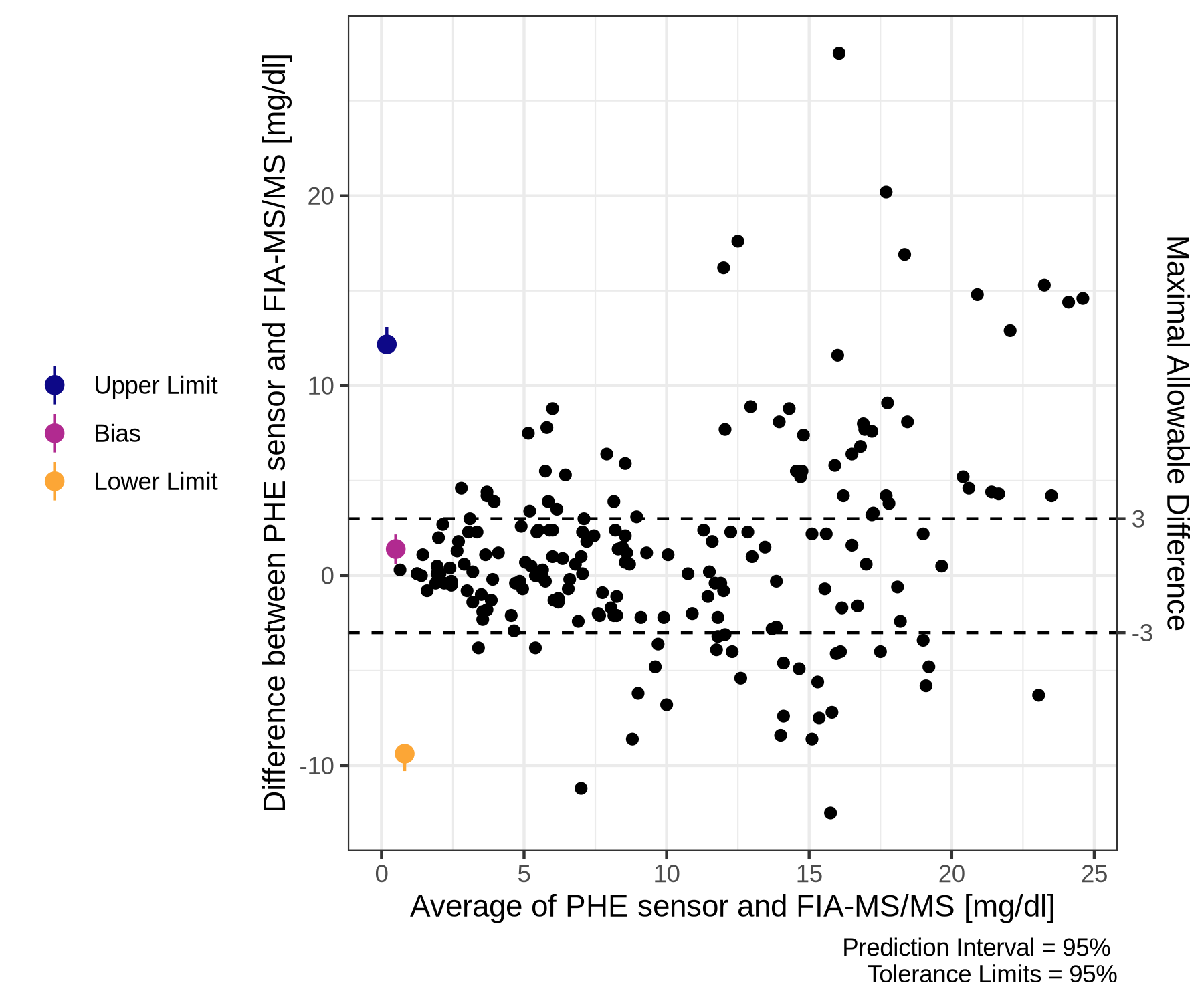

**Figure S2: Plot of tolerance.**

Scatter plot of differences between PHE sensor and FIA-MS/MS across average of both methods. The mean difference (bias), upper and lower tolerance limits with their confidence interval are indicated with yellow, purple and blue dots. The dashed horizontal lines indicate the upper limit of tolerated deviations set by the study physicians. The x-axis is labeled from 0 to 25 mg/dl PHE (0 to 1,500 µmol/l), and the y-axis from –10 to 20 mg/dl (–600 to 1,200 µmol/l).

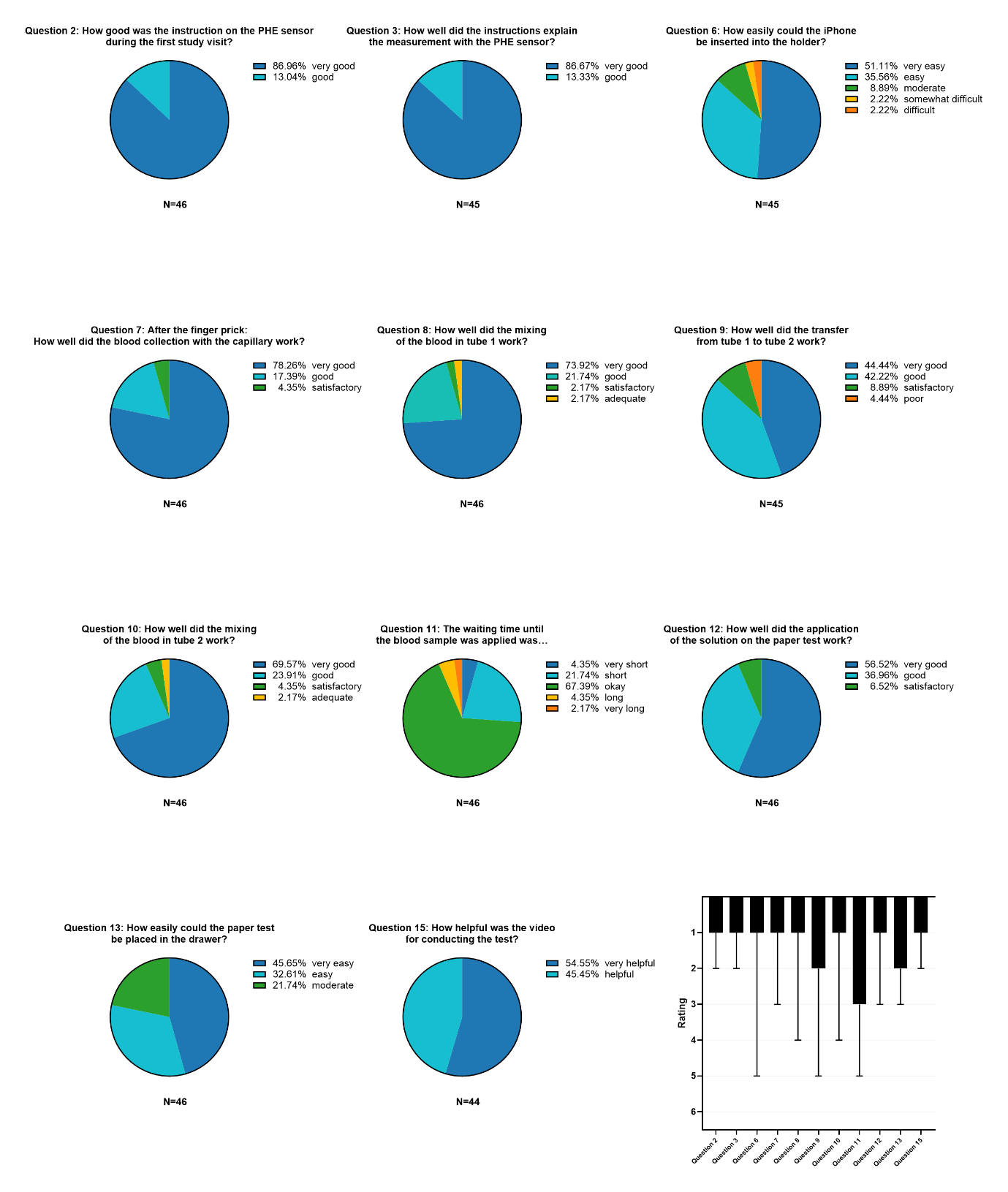

**Figure S3: Ratings of the instructions and test steps.**

Pie charts illustrate the percentage distribution of ratings; boxplots (scale 1 (best) to 6 (worst)) indicate the median, minimum (best rating), and maximum (worst rating) values.

**Table S2: Study schedule**

| **Study Visits** |  |  | **Screening** | **Visit 1** | **Visit 2** | **Visit 3** | **End of Study** |
| --- | --- | --- | --- | --- | --- | --- | --- |
| **Schedule: 0-12 years/ pregnant women** |  |  | **Day 1** | **2 weeks after screening (± 1 week)** | **4 weeks after screening (± 1 week)** | **6 weeks after screening (± 1 week)** | **8 weeks after screening (± 2 weeks)** |
| **Schedule: ≥ 13 years** |  |  | **Day 1** | **1 month after screening (± 1 week)** | **2 months after screening (± 1 week)** | **3 months after screening (± 1 week)** | **4 months after screening (± 2 weeks)** |
| Place of visit |  |  | Outpatient visit | At home (PoC) | At home (PoC) | At home (PoC) | At home (PoC) |
|  | Informed consent/ assent |  | X |  |  |  |  |
|  | Confirm eligibility |  | X |  |  |  |  |
|  | Enrollment |  | X |  |  |  |  |
|  | Vital signs, weight and height |  | X |  |  |  |  |
|  | Assessment of concomitant medications |  | X |  |  |  |  |
|  | PHE sensor levels |  | X | X | X | X | X |
|  | DBS PHE levels (FIA- MS/MS) |  | X | X | X | X | X |
|  | Questionnaire |  |  |  |  |  | X |
|  | Assessment of PHE sensor use related adverse events |  |  |  |  |  | X (via the questionnaire) |

**Table S3: Survey at the end of the study regarding the use of the PHE sensor and the *phenyx* app**

| 1 | Who performed the measurement with the PHE sensor? (Multiple answers possible) | Father of a PKU patient  Mother of a PKU patient  Both parents  Adult PKU patient  Adult female PKU patient (pregnant or not pregnant)  Boy with PKU  Girl with PKU  Someone else (please specify) |
| --- | --- | --- |
| 2 | How good was the instruction on the PHE sensor during the first study visit? | Very good  Good  Satisfactory  Adequate  Poor  Very poor  I don’t know / I prefer not to answer |
| 3 | How well did the instructions explain the measurement with the PHE sensor? | Very good  Good  Satisfactory  Adequate  Poor  Very poor  I don’t know / I prefer not to answer |
| 4 | How confident did you feel during the measurement? | Very confident  Confident  Mostly confident  Okay  Not confident  Not at all confident  I don’t know / I prefer not to answer |
| 5 | How long did it take for you to feel confident? | From the first test  From the second test  From the third test  From the fourth test  I still don't feel confident because …  I don’t know / I prefer not to answer |
| 6 | How easily could the iPhone be inserted into the holder? | Very easy  Easy  Moderate  Somewhat difficult  Difficult  Very difficult  I don’t know / I prefer not to answer |
| 7 | After the finger prick: How well did the blood collection with the capillary work? | Very good  Good  Satisfactory  Adequate  Poor  Very poor  I don’t know / I prefer not to answer |
| 8 | How well did the mixing of the blood in tube 1 work? | Very good  Good  Satisfactory  Adequate  Poor  Very poor  I don’t know / I prefer not to answer |
| 9 | How well did the transfer from tube 1 to tube 2 work? | Very good  Good  Satisfactory  Adequate  Poor  Very poor  I don’t know / I prefer not to answer |
| 10 | How well did the mixing of the blood in tube 2 work? | Very good  Good  Satisfactory  Adequate  Poor  Very poor  I don’t know / I prefer not to answer |
| 11 | The waiting time until the blood sample was applied was... | Very short  Short  Okay  Long  Very long  Too long  I don’t know / I prefer not to answer |
| 12 | How well did the application of the solution on the paper test work? | Very good  Good  Satisfactory  Adequate  Poor  Very poor  I don’t know / I prefer not to answer |
| 13 | How easily could the paper test be placed in the drawer? | Very easy  Easy  Moderate  Somewhat difficult  Difficult  Very difficult  I don’t know / I prefer not to answer |
| 14 | How would you rate the design of the *phenyx* app? | Very good  Good  Satisfactory  Adequate  Poor  Very poor  I don’t know / I prefer not to answer |
| 15 | How helpful was the video for conducting the test? | Very helpful  Helpful  Okay  Somewhat helpful  Not helpful  Not helpful at all  I don’t know / I prefer not to answer |
| 16 | How would you rate the overall handling of the PHE sensor? | Very good  Good  Satisfactory  Adequate  Poor  Very poor  I don’t know / I prefer not to answer |
| 17 | Did you experience any technical issues with the PHE sensor or the *phenyx* app? (Multiple answers possible) | No  Yes, finger prick using the lancet didn’t work  Yes, blood collection with the capillary didn’t work  Yes, combining blood and test solution didn’t work  Yes, applying the sample to the test paper didn’t work  Yes, inserting the test into the holder didn’t work  Yes, PHE measurement using the app didn’t work  Yes, the phenyx app didn’t work  Yes, ...  I don’t know / I prefer not to answer |
| 18 | If you had technical issues: How was the problem resolved? (Multiple answers possible) | I had to perform another finger prick  I dropped the remaining blood from tube 2 onto the measurement plate and measured again  I restarted the app  The problem could not be resolved  I don’t know / I prefer not to answer  Other solution, which... |
| 19 | How would you rate the overall effort with the PHE sensor and the *phenyx* app compared to conventional testing with the DBS? | Much lower  Lower  Similar  The same  Higher  Much higher  I don’t know / I prefer not to answer |
| 20 | Would you prefer to use the PHE sensor instead of DBS testing in the metabolic laboratory if it were available? | Yes  No |
| 21 | Would you recommend the PHE sensor to another patient with PKU? | Yes  No |
| 22 | What additional features would you like to see in the *phenyx* app? | … |
| 23 | Would you like to add anything or provide comments? | … |

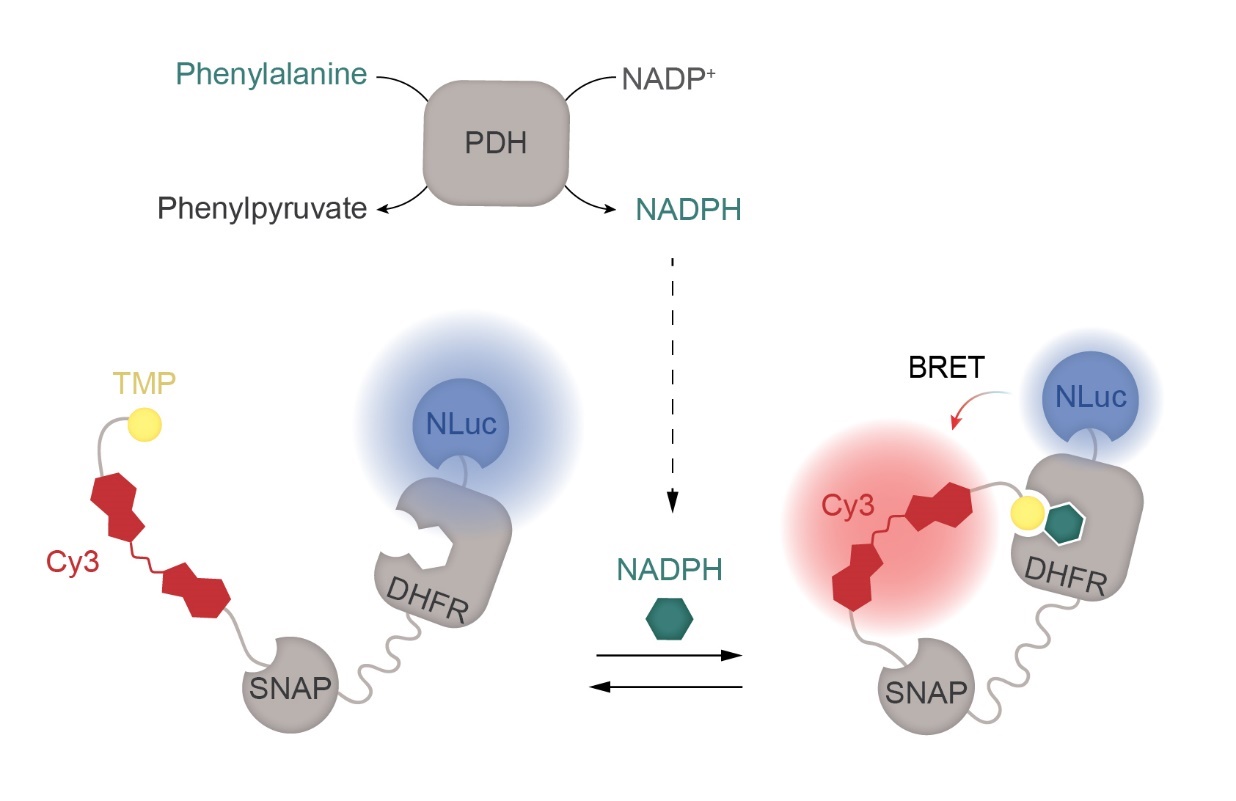

**Figure S4: PHE measurement principle using the NADPH sensor.**

The NADPH sensor is a bioluminescent fusion protein composed of a circularly permuted NanoLuc luciferase (Nluc), an NADPH-specific receptor dihydrofolate reductase (DHFR) and a self-labeling protein, SNAP-tag (SNAP). SNAP is labelled with a synthetic ligand containing a red Cyanine 3 (Cy3) fluorophore, and a DHFR inhibitor, trimethoprim derivative (TMP). TMP binds DHFR in the presence of NADPH, bringing Cy3 in close proximity to Nluc, resulting in bioluminescent resonance energy transfer (BRET) and the emission of red light. In the absence of NADPH, TMP does not bind DHFR resulting in the emission of blue light from Nluc. The ratio of emission intensities between Nluc and Cy3 is converted into NADPH concentration using a calibration curve. When paired with an NADP^+^-specific phenylalanine dehydrogenase (PDH), which converts PHE into phenylpyruvate and generates NADPH, the NADPH sensor allows for the quantification of PHE. Figure adapted from Yu *et al.*.^1^

**Detailed description of the PHE sensor kit and calibration**

The kit includes a lancet, two single-use pipettes, a tube containing buffer (tube 1), a tube with lyophilized reagents (tube 2), a test paper with four detection zones, and custom-designed holders for the tubes and the smartphone running the *phenyx* app. Lancet, pipettes, tubes 1 and 2 and test paper were stored with alcohol swab, plaster and dry silica pouch in a sealable aluminium pouch.

Tubes and single-use pipettes were purchased from Immundiagnostik AG (Bensheim, Germany; reference DZ1072). Tube 1 was filled and sealed by Immundiagnostik AG. Lancets were purchased from BD Microtainer (Heidelberg, Germany; reference 366593). Alcohol swabs were purchased from Ampri (Winsen, Germany; reference 09290). Plasters were purchased from Leukoplast (Hamburg, Germany; reference 79297-03). Dry silica pouches were purchased from Sumedtec. White sealable aluminium pouches were purchased from WACCOMT Pack. Aluminium adhesive foil was purchased from VWR (Radnor, USA; reference 606941-076) Test paper was purchased from Ahlström (Helsinki, Finland; reference 141). One-sided adhesive paper was purchased from TownStix (London, UK). Vivazine was purchased from Promega (Madison, USA; reference N2582). The other reagents were purchased from Sigma-Aldrich (Saint-Louis, USA).

Tube 1 is designed to lyse red blood cells within seconds, dilute the blood sample a 100-fold, and enable rapid and quantitative conversion of PHE to reduced nicotinamide adenine dinucleotide phosphate (NADPH) in Tube 2. Each tube was filled with 580 μL of 100 mM Glycine-KOH-KCl buffer (pH 10) supplemented with surfactants and sodium azide as a preservative.

Tube 2 is designed to convert PHE to NADPH and store the bioluminescent NADPH sensor responsible for signal transduction. In an empty tube, 500 μL of solution containing 200 nM PDH, 20 nM NADPH sensor, 4 mM NADP^+^, 1 U/mL esterase and excipients in 50 mM HEPES, 50 mM NaCl pH 7.13 was added. The tubes were first placed at -80°C for 2.5 hours before being transferred at -20°C for 2 hours, followed by an additional 2.5 hours at -80°C. After freezing, the tubes were placed into the lyophilizer (Alpha 2-4 LSCplus) for overnight lyophilisation at 0.1 mbar. Following the lyophilisation, tube 2 was manually sealed with aluminium adhesive foil.

The expression and purification of the NADPH sensor protein and PDH from *E. coli* strain BL21 (DE3) (Novagen), as well as the labeling of the NADPH sensor protein, were performed according to published procedures ^1^.

The paper test is designed to trigger bioluminescence from the NADPH sensor and enable PHE sensor measurement by the phenyx app. Test paper and one-sided adhesive paper were laser cut and adhesive paper folded around test paper, designing a strip featuring 4 circular detection zones. On 3 zones were spotted 5 µl of 1:50 Vivazine diluted in cold methanol. On the last control zone was spotted 5 µl of 1:50 Vivazine diluted in cold methanol supplemented with 1 mM PHE.

Custom-designed holders for the tubes and the smartphone were designed using Autodesk software and 3D printed (Ultimaker S5) by the Mechanical workshop at the Max-Planck Institute for Medical Research.

**Detailed instruction of use**

**Components:**

- Lancet
- 2x pipette
- Tube 1 and 2
- Paper test
- Smartphone holder with drawer
- iPhone with *phenyx* app
- Tube holder
- Dried blood card
- Disinfectant wipe
- Plaster
- Swab

**Step-by-step implementation of the test:**

1. Prepare iPhone
   - Ensure that the iPhone is sufficiently charged and connected to the internet.
   - Start the *phenyx* app.
   - Navigate through the app using the arrows and follow the instructions.
2. Scan QR code
   - Use the arrow to get to the "scan QR Code" screen.
   - Scan the QR code on the test packaging.
3. Open the test packaging
4. Place the iPhone in the phone holder
5. Place the tubes:
   - Place tubes 1 and 2 in the tube holder.
6. Pierce the foil:
   - Pierce the cover foil of tube 1 and tube 2 with the cap of a pipette.
7. Disinfect:
   - Clean the finger with a disinfectant wipe.
   - For infants: disinfect the heel.
   - It is recommended to use a warm environment, as the fingers have better blood circulation.
8. Prick your finger:
   - To take a blood sample with the lancet, remove the cap from the lancet and prick the prepared area on the fingertip (infants: heel).
9. Wipe away drops of blood:
   - Wipe away the first drop of blood with a swab (not the disinfectant wipe!).
10. Massage finger:
    - Massage your finger in the direction of the fingertip so that more blood comes out.
11. Carry out a dried blood spot test:
    - Fill one circle of the dry blood spot card up to the edge with blood.
    - Then put the card away.
12. Collect blood for paper test:
    - Place the tip of a disposable pipette vertically on the drop of blood until the pipette is full.
    - If air bubbles are visible, the process must be repeated with a new pipette from the emergency kit!
13. Transfer blood to tube 1:
    - Press the pipette filled with blood firmly into tube 1 until it clicks into place.
14. Shake tube 1:
    - Shake tube 1 together with the pipette for 10 seconds.
15. Stick on the plaster:
    - Apply a plaster to the puncture site.
16. Remove the cap:
    - Remove the cap from tube 1
17. Transfer the solution to tube 2:
    - Press tube 1 into tube 2 until it clicks into place.
    - Transfer the solution.
18. Shake tube 2:
    - Close tube 2 tightly with the second pipette until it clicks into place.
    - Shake the tube well for 10 seconds.
19. Waiting time:
    - Press "start waiting time".
    - Wait 5 minutes.
    - The total incubation time must not exceed 10 minutes! Once this time has elapsed, the test is no longer valid.
20. Remove the cap:
    - Remove the pipette cap from tube 2.
21. Insert paper test:
    - Place the paper test in the drawer of the smartphone holder.
22. Drop solution onto the paper test:
    - Hold tube 2 about 1 cm above the paper test.
    - Add one drop of the mixture to each of the 4 tests circle of the paper test.
23. Push the drawer in the smartphone holder:
    - Slide the drawer with the paper test in the smartphone holder.
    - Press the “start recording” button.
24. Leave the smartphone in the holder!
    - Pictures are now taken and sent automatically. Leave the smartphone until the timer has expired.
25. Disposal:
    - Dispose of all test components in the waste.

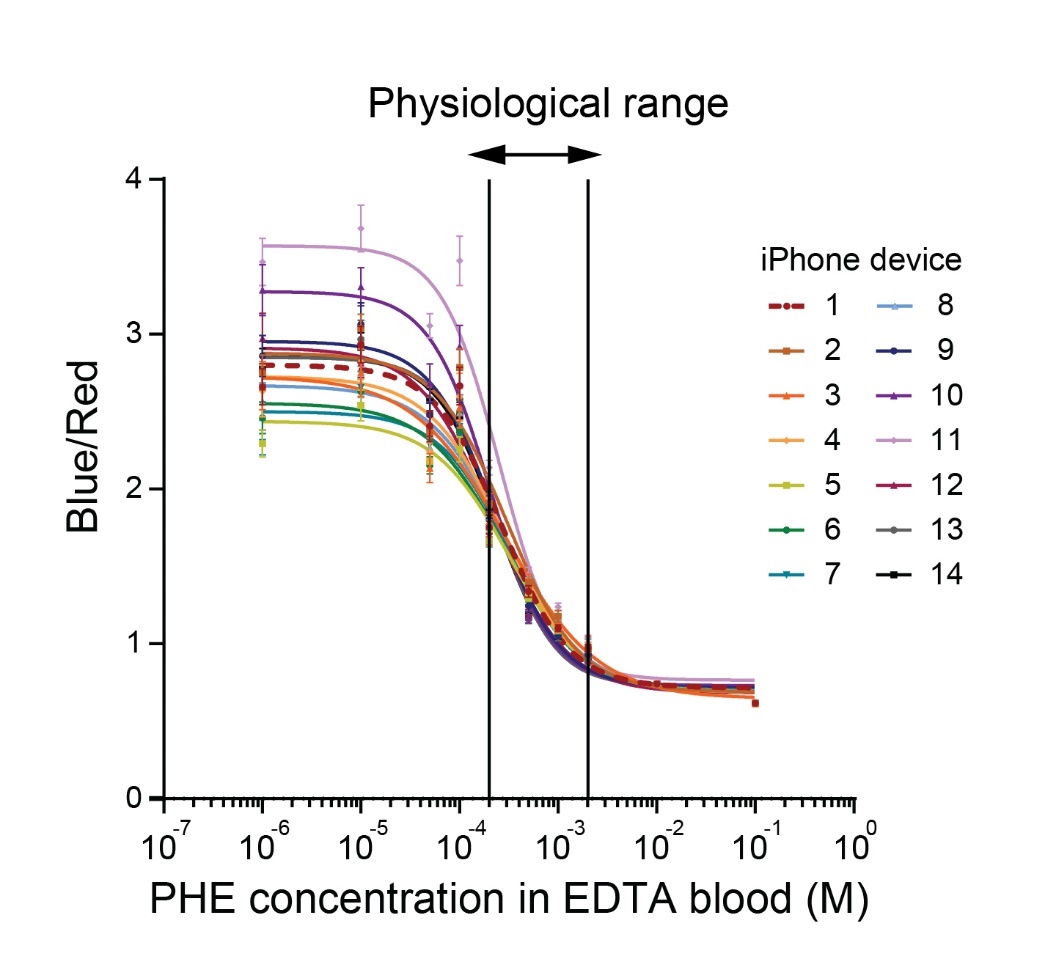

**Figure S5: Iphone device comparison.**

PHE sensor responses (Blue/Red ratios) were measured across 14 iPhone 14 Pro devices using the PHE sensor test kit with whole blood samples spiked with varying PHE concentrations. Šídák’s multiple comparisons test identified smartphones n°11 (p = 0.0017), n°10 (p < 0.0001), and n°5 (p < 0.0001) as significantly different from the reference device (smartphone n°1); these outlier devices were returned for replacement. The physiological range corresponds to the expected PHE concentration in blood in PKU patients.

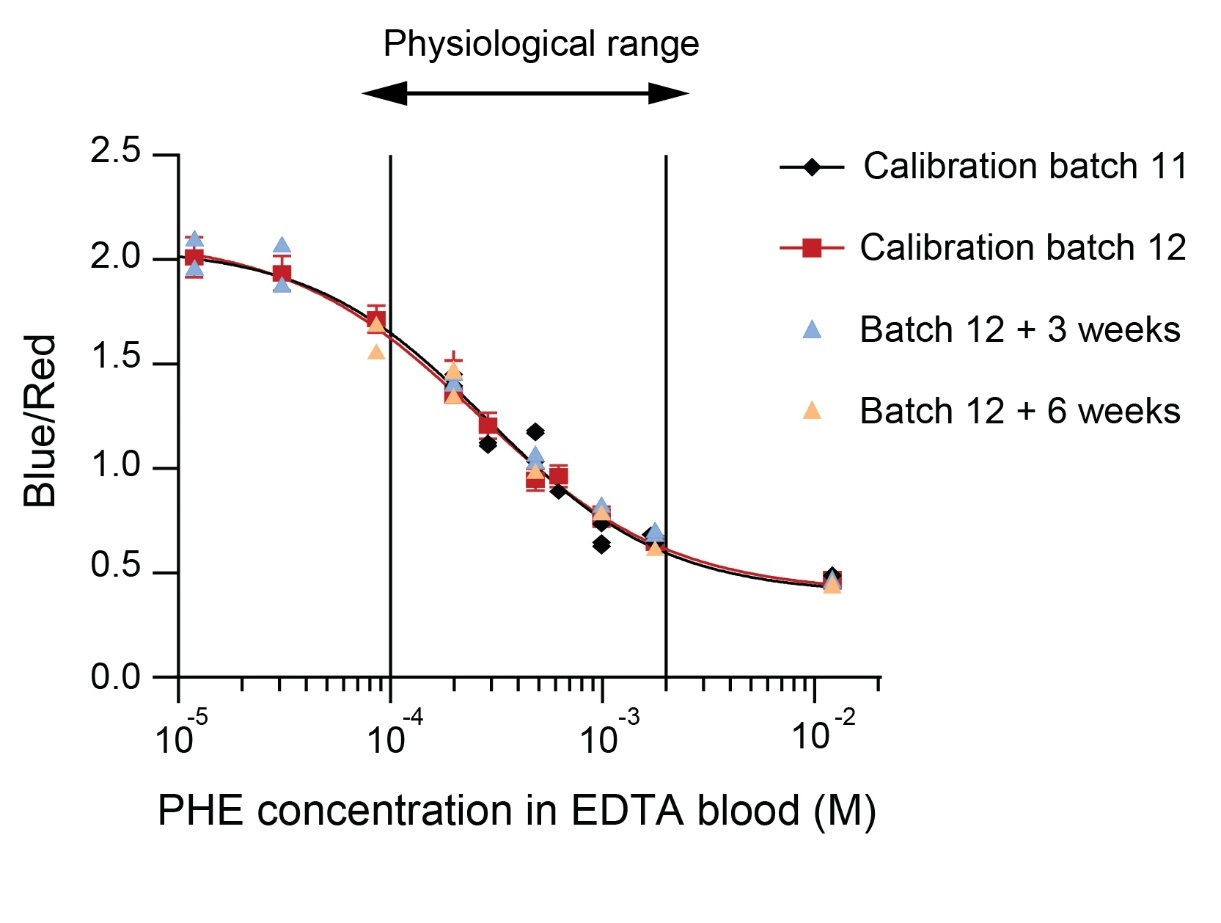

**Figure S6: PHE sensor calibration and inter-batch variation.**

Evaluation of PHE sensor performance including batch variation and stability after a 6-week storage period. The physiological range corresponds to the expected PHE concentration in blood in PKU patients.

**References**

1.	Yu, Q.*, et al.* Semisynthetic sensor proteins enable metabolic assays at the point of care. *Science* **361**, 1122-1126 (2018).
